# Genome-wide association study of Buruli ulcer in rural Benin

**DOI:** 10.1101/19012096

**Authors:** Jeremy Manry, Quentin B. Vincent, Maya Chrabieh, Lazaro Lorenzo, Ioannis Theodorou, Marie-Françoise Ardant, Christian Johnson, Estelle Marion, Annick Chauty, Laurent Marsollier, Laurent Abel, Alexandre Alcaïs, on behalf of the Franco-Beninese Buruli Research Group

## Abstract

Buruli ulcer, caused by *Mycobacterium ulcerans*, is the third mycobacterial disease worldwide characterized by devastating necrotizing skin lesions. The role of host genetics in susceptibility to Buruli ulcer has long been suggested. We conduct the first genome-wide association study of Buruli ulcer on a combined sample of 1,524 well characterized patients and controls from rural Benin. Two-stage analyses identify two novel associated loci located within lincRNA genes: rs9814705 in *ENSG00000240095*.*1* (*P* = 2.85×10^−7^; odds ratio = 1.80 [1.43-2.27]), and rs76647377 in *LINC01622* (*P* = 9.85×10^−8^; hazard ratio = 0.41 [0.28-0.60]). Furthermore, we replicate the protective effect of allele G of a missense variant located in *ATG16L1*, and previously shown to decrease bacterial autophagy (rs2241880, *P* = 0.003; odds ratio = 0.31 [0.14-0.68]). Our results suggest lincRNAs and the autophagy pathway as critical factors in the development of Buruli ulcer.

## Introduction

Buruli ulcer is a chronic infectious disease caused by *Mycobacterium ulcerans*. It received little attention until about 15 years ago, despite being more prevalent than tuberculosis or leprosy in some areas ^1^. Interest in this disease was stimulated by the identification, by the World Health Organization (WHO), of Buruli ulcer as a neglected emerging tropical disease and the launch of the Global Buruli Ulcer Initiative in 1998 ^2^. Buruli ulcer is now recognized as the third most frequent human mycobacterial disease worldwide, after tuberculosis and leprosy ^3^. It occurs mostly in rural areas of tropical countries, and West Africa is considered the principal endemic zone ^4^. Prevalence estimates between 2007 and 2016 ranged from 0.32 per 1,000 [95% confidence interval (CI) 0.31-0.33] in Ivory Coast to 2.99 per 1,000 [2.35-3.07] in Benin ^5^. In 2018, the WHO reported an increase in new cases of 39% relative to 2016 ^4^.

Buruli ulcer is a devastating necrotizing skin infection characterized by pre-ulcerative lesions (nodules, plaques, edema) that may develop into deep ulcers with undermined edges potentially extending to bones and joints. *M. ulcerans* induces painless skin necrosis through the production of mycolactone, a toxin that has been shown to play a key role in bacterial virulence and analgesia ^6, 7, 8, 9^. The painless nature of the disease often delays diagnosis, leading to late treatment and permanent disabilities, which affect more than 20% of patients, mostly children ^10^. Considerable variability has been reported in the clinical presentation of Buruli ulcer, including spontaneous healing in both humans ^11, 12^ and specific mouse strains ^12^. This observation, together with the familial clustering of cases ^13, 14^, supports the view of a substantial contribution of host genetic factors to the response to infection with *M. ulcerans*.

This hypothesis is consistent with the discovery of rare inborn errors of immunity conferring a predisposition to severe infections with weakly virulent mycobacteria, such as the BCG vaccine, in the context of the syndrome of Mendelian susceptibility to mycobacterial diseases (MSMD) ^15, 16^. Rare genetic defects have also been shown to underlie severe forms of tuberculosis ^17, 18^, and homozygosity for a missense polymorphism of *TYK2* was recently identified as the first common monogenic cause of tuberculosis ^19, 20^. A similar monogenic contribution to Buruli ulcer is supported by the recent identification of a microdeletion on chromosome 8 in a familial form of severe Buruli ulcer ^21^. Genome-wide association studies (GWAS) have identified several common variants associated with leprosy ^22, 23, 24^, while this kind of variants seems to play only a limited role in tuberculosis ^25^. By contrast, the role of common host genetic variants in the development of Buruli ulcer has been investigated in only three candidate-gene studies. These studies explored a total of seven genes selected on the basis of their involvement in MSMD, tuberculosis or leprosy. Significant association with Buruli ulcer was reported for common variants of six of these genes: *SLC11A1* ^26^, *PRKN, NOD2*, and *ATG16L1* ^27^ and *iNOS* and *IFNG* ^28^. We performed a two-stage case-control GWAS, to investigate comprehensively the role of common variants in the development of Buruli ulcer, on a sample of 1,524 individuals recruited over the last 15 years at the Centre de Dépistage et de Traitement de la Lèpre et de l’Ulcère de Buruli (CDTLUB) of Pobè, Benin.

## Results

### Genome-wide analyses

GWAS was performed on the discovery sample of 402 Buruli ulcer cases (sex ratio: 0.79; median age: 11 years) and 401 exposed controls (sex ratio: 0.72; median age: 40 years) living in villages in the Ouémé and Plateau *départements* in which Buruli ulcer is endemic (Table 1; Supplementary Fig. 1). Principal component analysis on genotyped variants with a MAF > 0.05 revealed no evidence of population stratification in our sample (Supplementary Fig. 2a), as all our individuals clustered with those of the Yoruba population of Ibadan, Nigeria (YRI) and the Esan population of Nigeria (ESN) from the 1000 Genomes Project (Supplementary Fig. 2a). Following imputation, 10,014,109 high-quality autosomal variants were tested for association with two different phenotypic definitions of Buruli ulcer. We first used the binary case/control Buruli ulcer phenotype and considered the additive genetic model (see Methods). The results of the GWAS association tests are summarized in a Manhattan plot (Fig. 1a). No substantial deviation from expectations was observed, as shown by the genomic inflation factor (λ=1.027) on the corresponding quantile-quantile plot (qq plot) (Supplementary Fig. 2a). In total, 517 variants had *P* values < 5×10^−5^, including 10 with *P* values < 10^-6^. The strongest signal for association was obtained for an imputed SNP (rs7246288) located on chromosome 19 with a *P* value = 2.80×10^−7^, a MAF of 0.09 and the odds ratio (OR) of developing Buruli ulcer for TT vs. CT or CT vs. CC of 2.33 [1.62-3.34]. After LD pruning, focusing on variants fulfilling the replication criteria (see Methods), 51 variants were retained for replication tests (Supplementary Table 1).

**Table 1.** Clinical characteristics of the discovery and replication samples.

| **Covariate** |  | **Discovery sample** |  | **Replication sample** |  | **Combined** |  |
| --- | --- | --- | --- | --- | --- | --- | --- |
|  |  | **Cases** | **Controls** | **Cases** | **Controls** | **Cases** | **Controls** |
| **Sex** | **Male** | 177 (44.0) | 168 (41.9) | 198 (43.5) | 98 (41.1) | 375 (43.8) | 266 (41.6) |
|  | **Female** | 225 (56.0) | 233 (58.1) | 257 (56.5) | 140 (58.9) | 482 (56.2) | 373 (58.4) |
|  | **Total** | 402 | 401 | 455 | 238 | 857 | 639 |
| **Age <sup>a</sup>** | **Mean age (SD)** | 18.5 (16.7) | 39.8 (17.3) | 22.4 (17.3) | 26.1 (18.7) | 20.5 (17.1) | 34.6 (19.0) |
|  | **Median age** | 11 | 40 | 15 | 22.5 | 13 | 35 |

NOTE. All values are expressed as *n* (%) unless otherwise indicated. SD: standard deviation.
<sup>a</sup> Age is given in years and corresponds to the age at the time of recruitment for controls and to age at diagnosis for cases.

**Figure 1.**
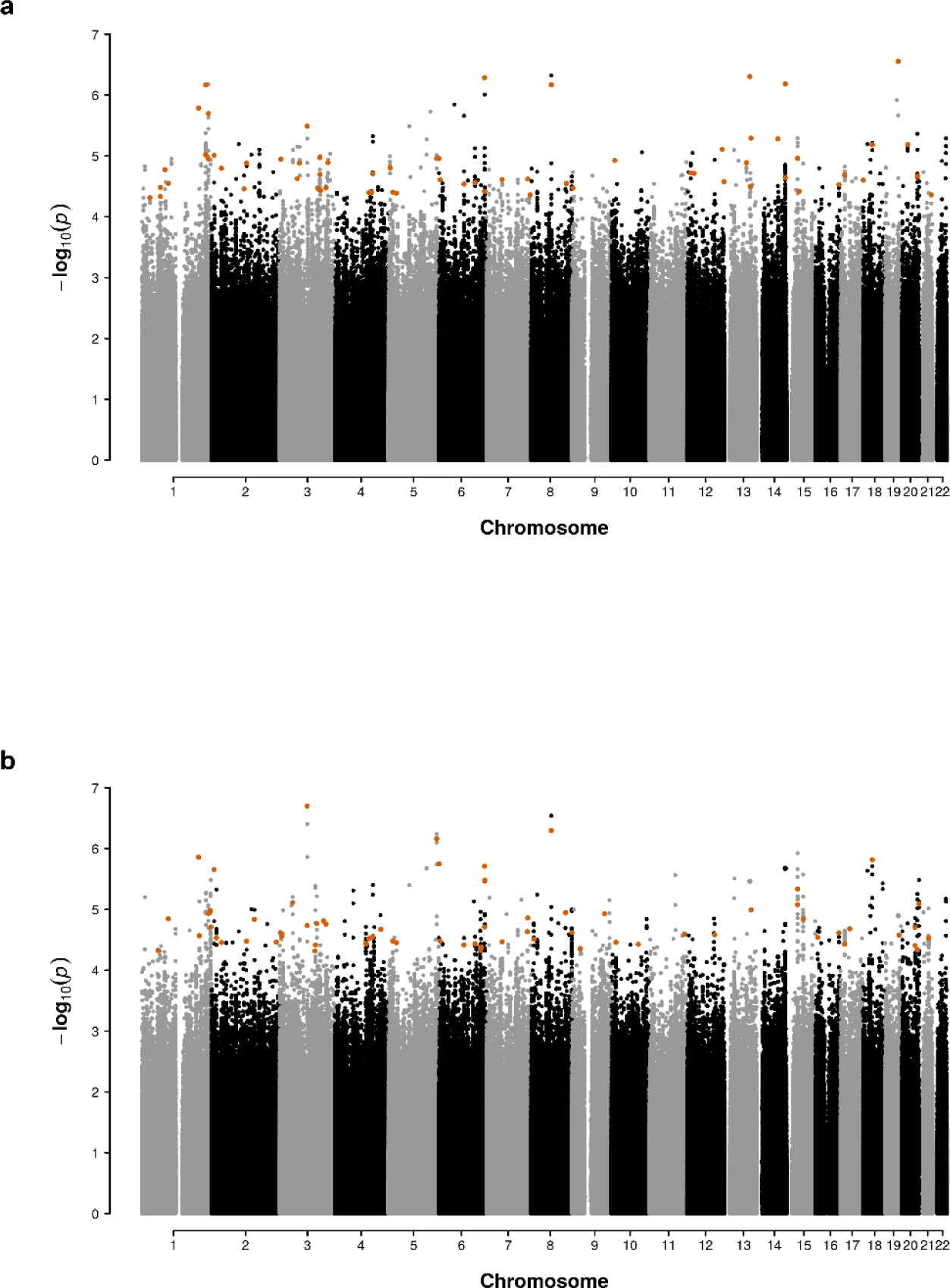
Manhattan plots of a GWAS of susceptibility to Buruli ulcer in a population from Benin. Manhattan plot of genome-wide *P* values for associations between variants and Buruli ulcer considering either a binary (affected/unaffected; panel A) or a censored (age at onset for Buruli ulcer patients and age at examination for the exposed controls; panel B) phenotype. The effects of variant genotypes on Buruli ulcer were investigated under an additive genetic model, using logistic regression and Cox models for the binary and censored phenotypes, respectively. The -log10*P*-values (*y-*axis) for SNPs are presented according to chromosomal positions (*x-*axis). Larger orange dots correspond to the 105 variants selected for replication.

Age at diagnosis can provide important information about the architecture of the genetic contribution to the disease. We performed another GWAS taking age at diagnosis of Buruli ulcer into account with a Cox model (see Methods). The corresponding qq plot (λ = 1.023) and Manhattan plot are shown in Supplementary Fig. 2a and Fig. 1b, respectively. In total, 534 variants had an association *P* value < 5×10^−5^, including nine with a *P* value < 10^-6^. The strongest association signal was obtained for a genotyped SNP (rs34060873) located on chromosome 3 (*P* value = 1.99×10^−7^). The minor allele A (MAF = 0.10) was protective, with a hazard ratio (HR) for the development of Buruli ulcer for AA *vs*. CA or CA *vs*. CC estimated at 0.51 [0.39-0.67]. The same LD pruning strategy led to the selection of 54 variants for replication analyses (Supplementary Table 1). Overall, considering the two phenotypic definitions described above as probably having independent signals, 105 variants, including 99 genotyped variants, were selected for replication analysis (Supplementary Table 1).

### Replication study

In the second phase, the 105 identified variants were genotyped in a replication sample composed of 455 cases (sex ratio: 0.77; median age: 15 years) and 238 controls (sex ratio: 0.70; median age: 22.5 years) (Table 1). After quality control, 100 variants were retained for association testing in the replication sample with the same phenotypic models (binary and age at onset) and the same allelic effect as observed for the discovery sample (Supplementary Table 1). Five variants could not be tested. Three of these variants were imputed in the discovery sample and could not be genotyped. Two were present in Alu sequences and the other in an LRT element, which are prone to genotyping and imputation errors. The two remaining variants were genotyped in the discovery sample but were found to be in Hardy-Weinberg disequilibrium among the controls of the replication sample (*P* value < 10^-4^) (Supplementary Table 1). For the 100 high-quality variants retained, evidence of replication, with a *P* value less than 0.01 in one-tailed tests, was obtained for two SNPs (Supplementary Table 1 and Table 2).

The first evidence of replication was observed for the binary phenotype and rs9814705 on chromosome 3, for which the replication *P* value was 6.5×10^−4^. As this variant had been imputed in the discovery sample, we decided to subject it to Sanger sequencing in the discovery sample. This resequencing effort slightly increased the discovery *P* value from 3.71×10^−5^ (Supplementary Table 1) to 1.67×10^−4^ (Table 2). The combined *P* value for this SNP in our two samples was close to the genome-wide significance level (combined *P* value = 2.85×10^−7^). The minor allele C was the risk allele (MAF = 0.14), and the OR for developing Buruli ulcer for CC *vs*. TC or TC *vs*. TT was estimated at 1.80 [95% CI, 1.43-2.27]) (Table 2). The proportion of Buruli ulcer patients was 53% for TT carriers, and 76% for CC carriers (Fig. 2, Table 3). We then looked for additional variants in strong LD (r^2^ > 0.5) with rs9814705, using the 1000 Genomes Phase III data for the Yoruba population. Three SNPs were identified: rs1513419 (r^2^ = 0.87, GWAS *P* value = 4.37×10^−5^), rs7637582 and rs7615284 (r^2^ = 0.69, and GWAS *P* value = 1.01×10^−5^ for both) (Fig. 3a). This cluster of four variants is located on chromosome 3 in an intron of a lincRNA (ENSG00000240095.1) of unknown function, with the closest protein-coding gene (*PLOD2*) 107 kb away.

**Table 2.**
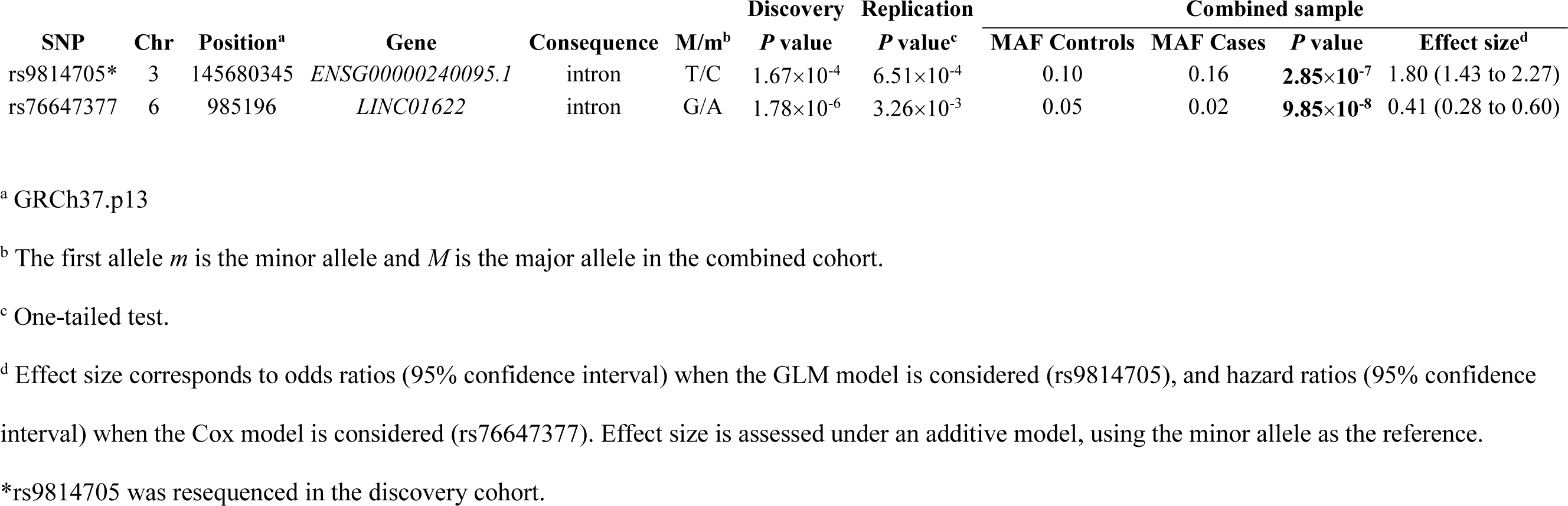
Association results for the two replicated Buruli ulcer GWAS signals.

| SNP | Chr | Position<sup>a</sup> | Gene | Consequence | M/m<sup>b</sup> | Discovery | Replication | Combined sample |  |  |  |
| --- | --- | --- | --- | --- | --- | --- | --- | --- | --- | --- | --- |
|  |  |  |  |  |  | *P* value | *P* value<sup>c</sup> | MAF Controls | MAF Cases | *P* value | Effect size<sup>d</sup> |
| rs9814705* | 3 | 145680345 | *ENSG00000240095.1* | intron | T/C | 1.67×10<sup>-4</sup> | 6.51×10<sup>-4</sup> | 0.10 | 0.16 | **2.85×10<sup>-7</sup>** | 1.80 (1.43 to 2.27) |
| rs76647377 | 6 | 985196 | *LINC01622* | intron | G/A | 1.78×10<sup>-6</sup> | 3.26×10<sup>-3</sup> | 0.05 | 0.02 | **9.85×10<sup>-8</sup>** | 0.41 (0.28 to 0.60) |

<sup>a</sup> GRCh37.p13
<sup>b</sup> The first allele *m* is the minor allele and *M* is the major allele in the combined cohort.
<sup>c</sup> One-tailed test.
<sup>d</sup> Effect size corresponds to odds ratios (95% confidence interval) when the GLM model is considered (rs9814705), and hazard ratios (95% confidence interval) when the Cox model is considered (rs76647377). Effect size is assessed under an additive model, using the minor allele as the reference.
\*rs9814705 was resequenced in the discovery cohort.

**Figure 2.**
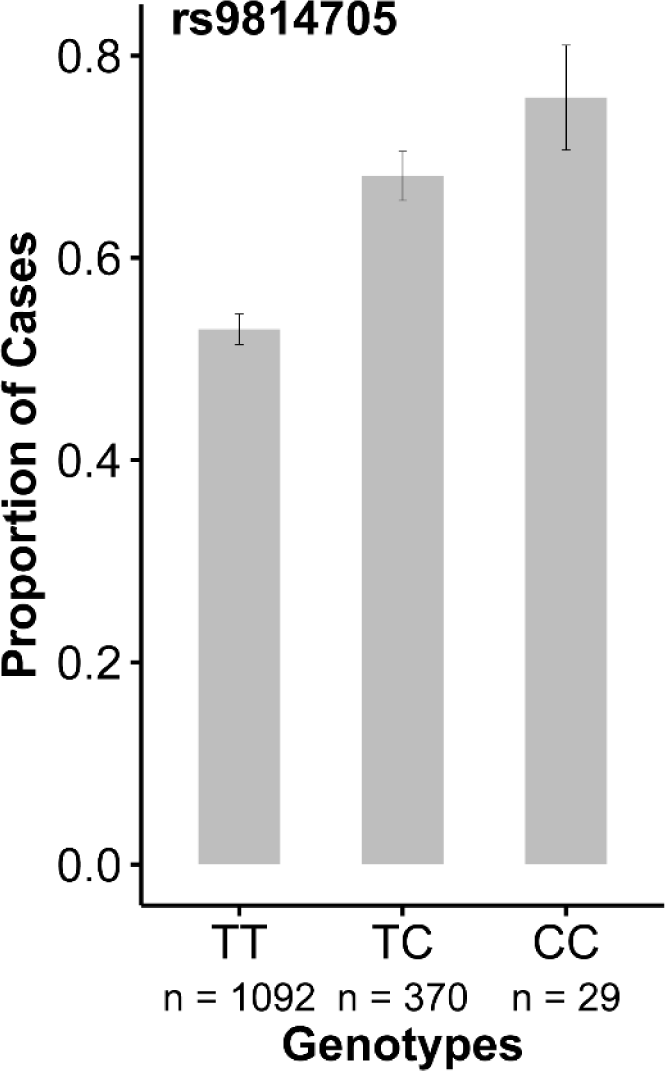
Genotype at rs9814705 is associated with case-control status. Distribution of the proportion of Buruli ulcer cases (*y*-axis) according to rs9814705 genotype (*x*-axis) in the combined sample (*n*: number of individuals).

**Figure 3.**
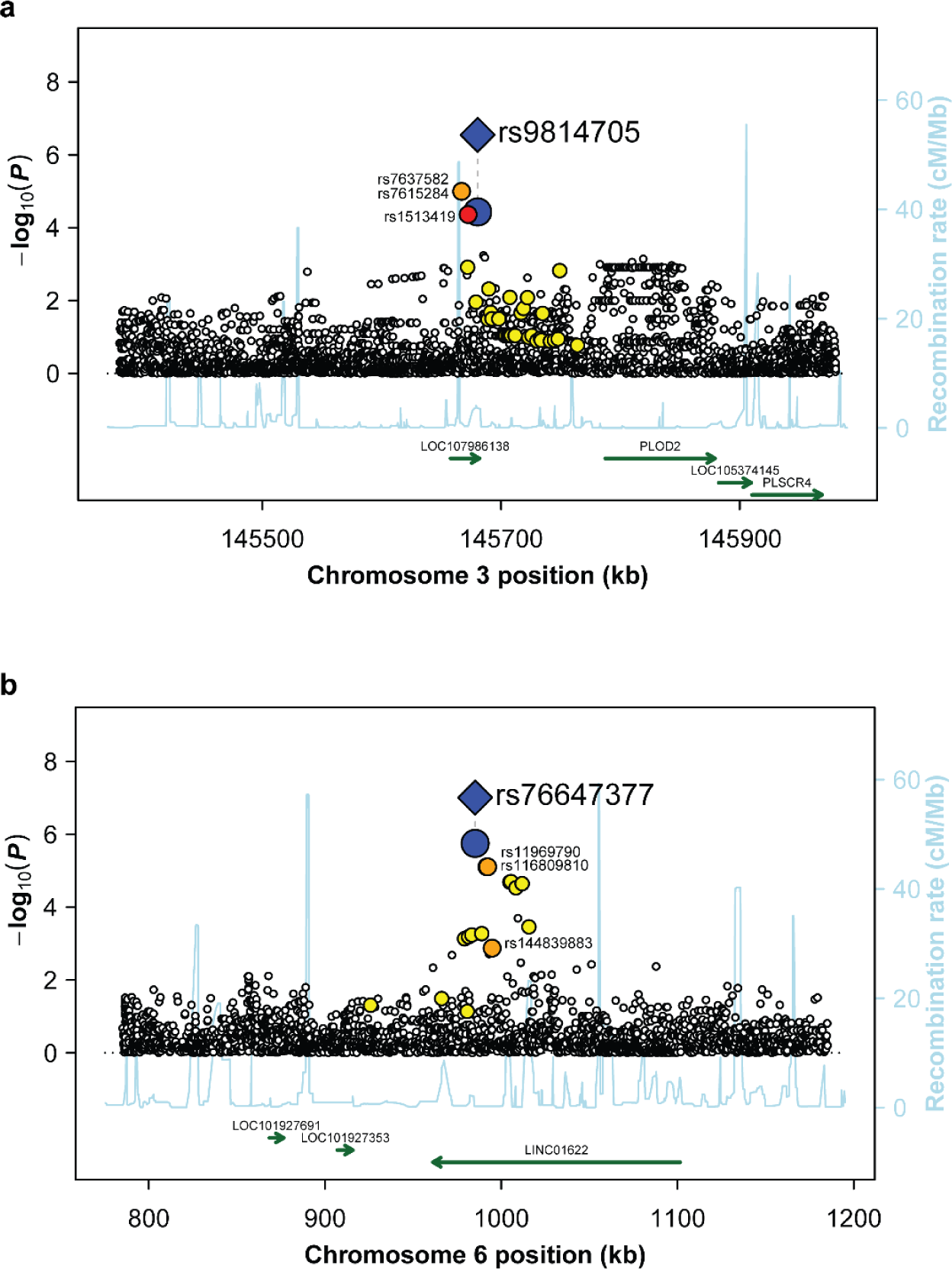
Association, linkage disequilibrium and gene maps for the two replicated association signals. Plots, with GRCh37 coordinates, showing evidence of association (-log10(*P*); left *y*-axis) for the variants located in the vicinity of (A) rs9814705 and (B) rs76647377 (kb; *x*-axis). The distribution of recombination rates in these regions is also given (cM/Mb; Right *y*-axis; light blue line). For the two replicated SNPs (i.e. rs9814705 and rs76647377), blue diamonds correspond to the results observed in the combined analysis, whereas blue circles correspond to the results observed in the GWAS only. A color-coded scheme is used to display the degree of LD of the proximal variants with the replicated SNP (red: r^2^ > 0.8, orange: 0.5 < r^2^ < 0.8, and yellow: 0.2 < r^2^ < 0.5). Only variants with an r^2^ > 0.5 are labeled. Genes in the two chromosomal regions are plotted, with arrows indicating their orientation.

**Table 3.** Genotypic distributions of the two replicated SNPs between cases and controls in the combined cohort.

| **SNP** | **Chr** | **Position<sup>a</sup>** | **Group** | **Genotype Frequencies (n)<sup>b</sup>** |  |  |
| --- | --- | --- | --- | --- | --- | --- |
| **rs9814705** | **3** | **145680345** |  | **T/T** | **T/C** | **C/C** |
|  |  |  | **Cases** | 0.678 (578) | 0.296 (252) | 0.026 (22) |
|  |  |  | **Controls** | 0.804 (514) | 0.185 (118) | 0.011 (7) |
| **rs76647377** | **6** | **985196** |  | **G/G** | **G/A** | **A/A** |
|  |  |  | **Cases** | 0.968 (827) | 0.032 (27) | 0 (0) |
|  |  |  | **Controls** | 0.909 (582) | 0.089 (57) | 0.002 (1) |

<sup>a</sup> GRCh37.p13
<sup>b</sup> Missing Sanger sequencing data for rs9814705 (143 samples) were obtained by best-guess imputation.

The second replicated SNP was rs76647377 on chromosome 6, which had an association *P* value of 3.26×10^−3^ in the second sample based on the age-of-onset phenotype. The combined *P* value for this SNP was close to the genome-wide significance level at 9.85×10^−8^. The minor allele A (MAF = 0.03) was protective, with a HR for developing Buruli ulcer for AA *vs*. GA or GA *vs*. GG estimated at 0.41 [0.28-0.60]) (Table 2). GG carriers were found to be prone to developing the disease at an earlier age than GA or AA carriers (Fig. 4). Given that there was only one AA individual, who was a control, we were unable to distinguish between an additive model and a dominant model for allele A, as the combineddo not have the power to discriminate between an additive model, and a dominant model for allele A that provided a very close combined *P* value at 1.37×10^−7^. We identified three additional variants with an r^2^ > 0.5 with rs76647377: rs116809810 and rs11969790 (r^2^ = 0.58, GWAS *P* value = 4.55×10^−5^ for both), and rs144839883 (r^2^ = 0.54, GWAS *P* value = 2.7×10^−3^) (Fig. 3b). All these variants are located on chromosome 6, in the intron of a lincRNA *LINC01622* of unknown function (Table 2), and the closest protein-coding gene is *EXOC2*, 292 kb away.

**Figure 4.**
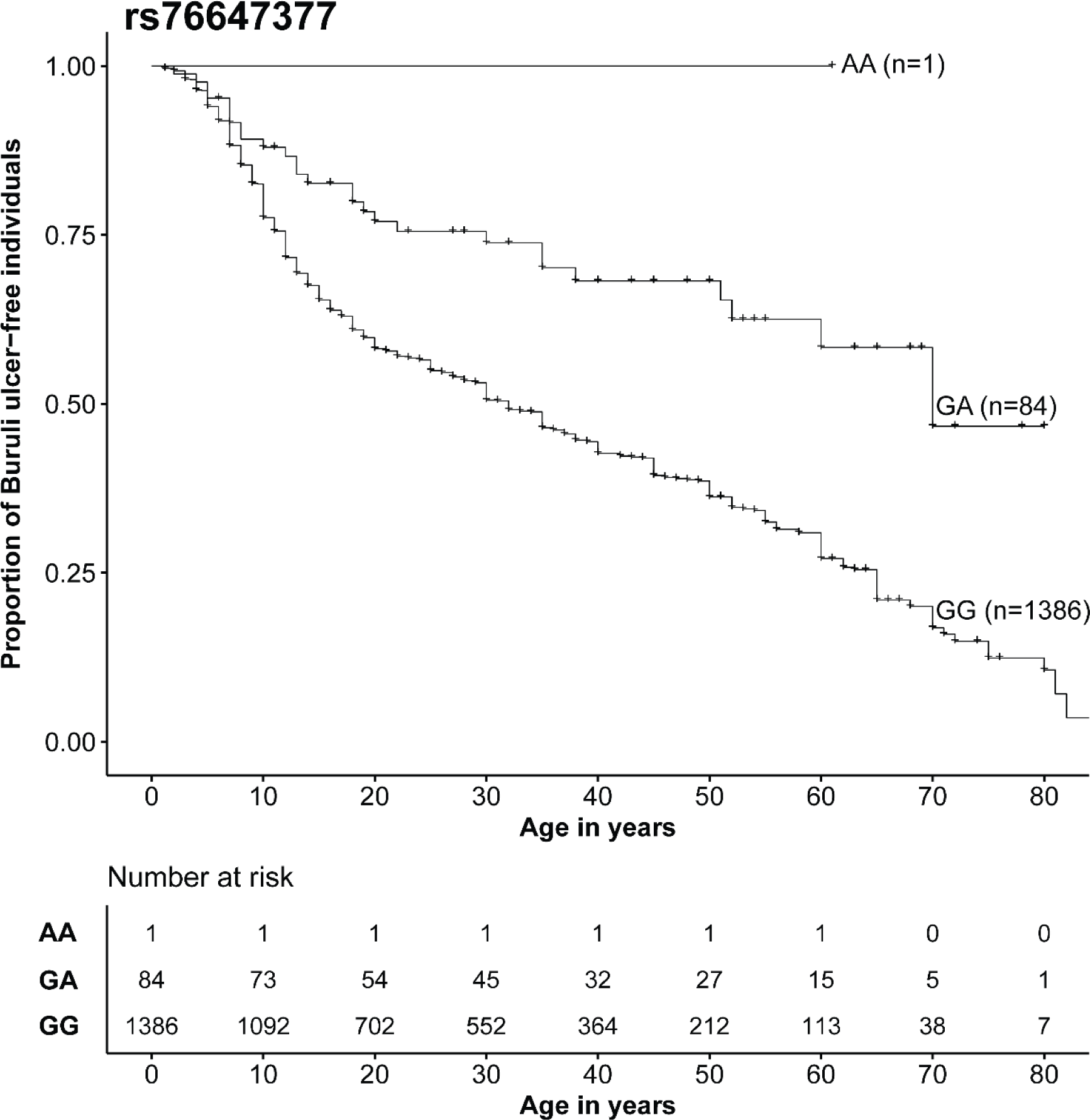
Genotype at rs76647377 is associated with age at onset of Buruli ulcer. Kaplan-Meier curves for the onset of Buruli ulcer (*y*-axis) according to rs76647377 genotype. Number of individuals at risk are provided by genotype and by ten-year age bins.

### Signals overlapping with previously reported associations

As the microdeletion recently identified in a familial form of Buruli ulcer was close to a cluster of beta-defensin genes ^21^, we also investigated the role of 5,447 variants located in 50 defensin genes and 91 eQTL for defensin genes (Supplementary Table 2). No enrichment in *P* values either < 0.01 or < 0.001 was observed among these variants in our discovery sample, the best hit being rs58925751, located in the *DEFB123* gene on chromosome 20 (*P* value = 4.1×10^−4^). We then checked the level of association in our GWAS for the six variants displaying significant association signals in the only three other candidate-gene association studies on Buruli ulcer performed to date ^26, 27, 28^ (Table 4). Only one of these variants — rs2241880, a missense T300A variant located in the *ATG16L1* gene — had a *P* value < 0.05 in our GWAS under the additive model with the same allelic effect (one-tailed *P* value = 0.015). The association was even more significant (*P* value = 0.009) when considering a recessive model for allele G, the model of inheritance reported in the original work. Under this model, the OR for developing Buruli ulcer for GG vs. GA or AA subjects was estimated at 0.46 [0.25-0.86]. Furthermore, in the previous study ^27^, GG homozygotes were found to be protected against the ulcerative clinical form of Buruli ulcer, with an OR of 0.35 [0.13–0.90]. Strikingly, when focusing only on ulcerative cases of our discovery sample (255 ulcerative cases *vs*. 401 controls), the association was more significant (*P* value = 0.003), with an OR of 0.31 [0.14-0.68] (Table 4).

**Table 4.**
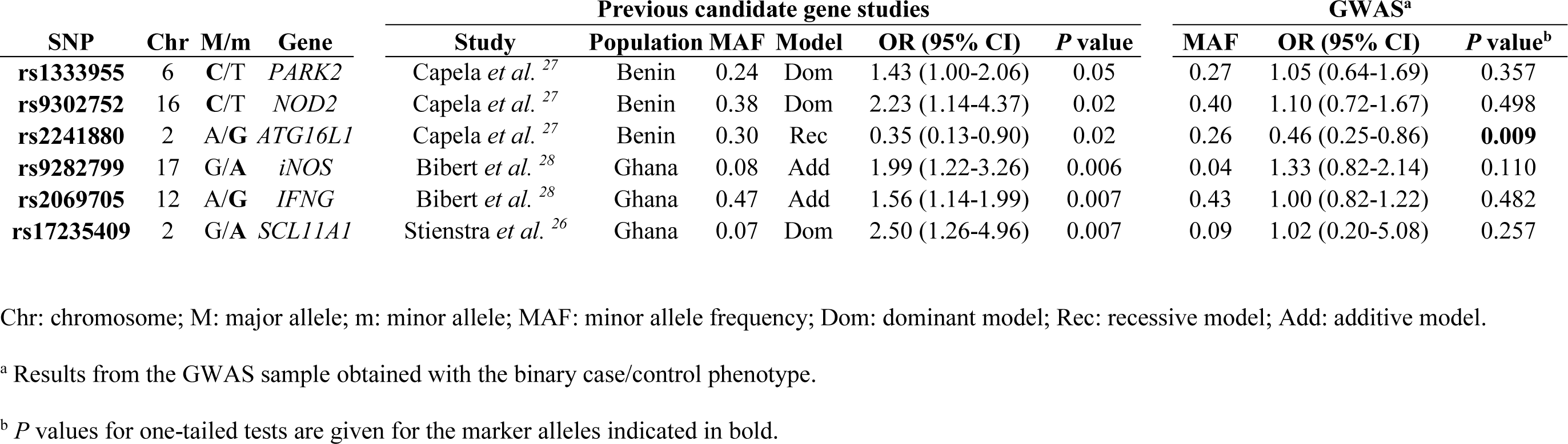
Validation study for six variants of three genes previously reported to be associated with Buruli ulcer.

Finally, we also investigated whether some SNPs found to be associated with the other two most common mycobacterial diseases, tuberculosis and leprosy, overlapped with the SNPs identified in our GWAS on Buruli ulcer (see Methods for details). We first looked at the 105 SNPs selected for replication in our GWAS, and noted that rs9295218 — the strongest hit among the genotyped SNPs identified in our discovery sample (GWAS *P* value = 5.17×10^−7^; OR = 1.64 [1.35-2]) — was located in an intron of the *PACRG* gene, variants of which have been associated with leprosy ^29^ (Supplementary Table 1). We observed the same trend towards association in the replication sample, although it was not significant (replication *P* value = 0.13), with an OR of 1.14 [0.91-1.42]). This variant is not in LD with variants of the *PRKN* (also called *PARK2*)*/PACRG* cluster associated with leprosy, including that reported to be associated with Buruli ulcer by Capela *et al*. ^27^ (r^2^ = 0.007) (Table 4). We then looked at the 35 SNPs shown to be of genome-wide significance in the GWAS performed on tuberculosis (eight independent variants; Supplementary Table 3a) or leprosy (27 variants in 19 different chromosomal regions; Supplementary Table 3b). None of these 35 SNPs was found to be associated with Buruli ulcer with a *P* value < 0.01. The lowest *P* value (0.017) was obtained for rs2269497, a missense variant of *RGS12* identified in a GWAS on tuberculosis in a Chinese population ^30^, for which the minor G allele was found to be associated with a risk of tuberculosis, whereas we found it to have a protective effect in Buruli ulcer. Taken together, these results suggest that there is no strong overlap between common variants potentially involved in the development of Buruli ulcer and in that of tuberculosis or leprosy.

## Discussion

Less than 3% of GWAS to date have focused on sub-Saharan African populations (*vs*. ∼50% on populations of European ancestry) ^31^. The inaccessibility of many areas makes it difficult to set up GWAS focusing on a neglected tropical disease. In this context, we report the first GWAS investigating susceptibility to Buruli ulcer in a well-characterized sample, including more than 1,500 individuals, in Benin. This sample may be smaller than those used in GWAS on more common infectious diseases, such as leprosy and tuberculosis, but it is the largest sample ever used for a genetic association study on Buruli ulcer. The discovery sample had a power of 80% for detecting, with a type I error of 5×10^−5^ (the threshold to be tested for replication), variants with a MAF > 0.18 and an OR >1.8. Individuals were recruited for this sample from areas of Benin in which Buruli ulcer is highly endemic, from the villages of Ouémé and Plateau (Supplementary Fig. 2a). The high local prevalence in these areas, and our study design, in which the controls were older than the cases, maximize the chances of controls having been exposed to *M. ulcerans*. The enrollment of unexposed controls would decrease the power of the study. In addition, the definition of Buruli ulcer for the discovery sample was based strictly on the laboratory confirmation of cases, increasing the quality of the criteria used for selecting cases and controls in this study.

The GWAS results highlighted two independent loci displaying a strongly suggestive association with Buruli ulcer rs9814705 for analyses with the binary affected status (*P* value = 2.85×10^−7^), and rs76647377 when age at Buruli ulcer onset was taken into account (*P* value = 9.85×10^−8^). These two variants are in regions of the genome devoid of protein-coding genes, the nearest such genes being *PLOD2* (107 kb away) for rs9814705, and *EXOC2* (292 kb away) for rs76647377. Both variants were located within the introns of lincRNAs: *ENSG00000240095*.*1* (rs9814705) and *LINC01622* (rs76647377). Unfortunately, few (for rs9814705) or no (for rs76647377) functional data are available for these lincRNAs, the functions of which remain unknown. SNPs rs9814705 and rs1513419 (r^2^ =0.87 with rs9814705) are eQTL for lincRNA *ENSG00000240095*.*1* in esophagus-mucosa tissue (*P* value = 3.9×10^−5^ and *P* value = 4.5×10^−5^, respectively) ^32^. In the GTEX data, this lincRNA appeared to be expressed only in esophagus-mucosa tissue and in the minor salivary gland. In addition, rs9814705 and rs1513419 had *P* values of borderline significance (0.049) for being cis-eQTL of *PLSCR1* in lipopolysaccharide-stimulated monocytes from healthy individuals ^33^. The *PLSCR1* gene, located 565 kb away from rs9814705, encodes the transmembrane protein phospholipid scramblase 1, which has been shown to be overexpressed after stimulation with type I interferons, and to play a crucial role in apoptosis ^34, 35^. This gene has also been identified as a core gene in the host response to tuberculosis in a meta-analysis of transcriptomic data ^36^. Thus, *PLSCR1* may be an interesting candidate gene for involvement in the development of Buruli ulcer. Further studies are required to determine the precise role of the identified lincRNAs.

Using our GWAS data, we replicated the effect of the A/G SNP rs2241880, located in the *ATG16L1* gene, initially identified in another sample from Benin ^27^. Individuals homozygous for the minor allele G (MAF of 0.33 in the African populations of gnomAD) are protected against Buruli ulcer, especially the ulcerative forms, with an OR of 0.31 [0.14-0.68] in our sample and 0.35 [0.13-0.90] in the original study. The protein encoded by *ATG16L1* is required for autophagy ^37^, and the G allele has been associated with an increase in the release of cytokines, such as IL-1β and IL-18, leading to a decrease in antibacterial autophagy ^38, 39^. This variant results in a T300A missense substitution in the principal isoform of *ATG16L1*. Remarkably, it has also been reported to be an eQTL in nine tissues, including skin, in which the G allele of *ATG16L1* is associated with higher levels of expression of this gene (*P* value = 1.6×10^−25^) ^32^. Allele G of rs2241880 was also found to increase the risk of Crohn’s disease (CD) in several GWAS, with an OR of about 1.4 under an additive model ^40, 41, 42^. In addition, *Atg16L1*-hypomorphic mice developed intestinal abnormalities resembling CD whilst displaying resistance to intestinal infection with the bacterium *Citrobacter rodentium* ^43, 44^. Published findings suggest that autophagy may serve as a rheostat for immune reactions ^45^. These opposite effects in Buruli ulcer and CD provide another example of mirror genetic effects (*i*.*e*. an allele protective against infection but deleterious for inflammatory disorders), consistent with the view that the current high frequency of inflammatory/autoimmune diseases may reflect past selection for strong immune responses to combat infection ^46^.

Despite significant practical and methodological challenges ^47, 48, 49^, we successfully conducted a GWAS on a neglected tropical disease in rural sub-Saharan Africa, discovered two loci associated with Buruli ulcer and replicated a previously identified locus. Further studies are required to decipher the role of these variants and the function of the lincRNAs in which they are located. Detailed transcriptomic and epigenetic profiles for both whole blood (systemic response) and at the site of ulcers (local response) would complement our genomic approach and highlight genes and pathways involved in the disease. Mouse models of Buruli ulcer ^50^ will also be of major interest in this context, for further investigation of the role of the *ATG16L1* variant in the response to *M. ulcerans*, in particular.

## Methods

### Ethics statement

This study on genetic susceptibility to Buruli ulcer was approved by the institutional review board of the CDTLUB (Pobè, Benin), the National Ethical Review Board of the Ministry of Health in Benin (IRB00006860), and the ethics committee of the University Hospital of Angers, France (Comité d’Ethique du CHU d’Angers). Written informed consent was obtained from all adult participants.Parents or guardians provided informed consent on behalf of all minors participating in the study.

### Subjects

Between July 2003 and June 2012, 1,524 individuals from the CDTLUB in Pobè, Benin, were enrolled. All participants were living in villages distributed over the Ouémé and Plateau *départements* (an administrative area equivalent to a county) in an area in which Buruli ulcer was endemic ^51, 52^. Based on the cases registered in these two areas between 2005 and 2012, the overall prevalence in the studied villages was 5.3 per 1,000; a detailed distribution by village of the cases recorded is shown in Supplementary Fig. 2a. This discovery sample consisted of 408 HIV-free Buruli ulcer cases and 408 exposed controls (354 male and 462 female subjects; the mean ages of the cases and controls were 18.5 and 39.8 years, respectively). We chose to include controls that were older than the cases, as being older meant that they had been exposed to *M. ulcerans* for a longer time but had remained Buruli ulcer-free). All 408 cases were confirmed by IS2404 PCR, culture and/or Ziehl-Neelsen staining in the laboratory ^10, 53^. The replication sample consisted of 708 individuals: 467 HIV-free Buruli ulcer patients and 241 exposed controls (303 male and 405 female subjects; the mean ages of cases and controls were 22.3 and 26.1 years, respectively). The cases included in the replication sample were either laboratory-confirmed (48.6%) or classified as highly probable (51.4%) on the basis of stringent clinical criteria and treatment efficacy.

### Genotyping, imputation and quality control

Genomic DNA was extracted from 5 to 10 mL blood samples with the Nucleon BACC2 Genomic DNA extraction kit (GE Healthcare), in accordance with the manufacturer’s instructions. DNA preparations from the discovery sample were randomized in nine 96-well plates (with an equal number of cases and controls per plate) and genotyped with the Illumina Omni2.5 chip array. Genotypes were assessed with GenTrain implemented in Illumina GenomeStudio software (v2011.1). Thirteen of the 816 samples were excluded from further analyses due to a call rate <95% (8 samples), lack of concordance between reported and genetically inferred gender (3 samples), or duplication (2 samples). Our ‘effective’ discovery sample therefore consisted of a total of 803 individuals: 402 cases and 401 controls (Table 1).

In total, we obtained 2,314,174 non-filtered autosomal variants, of which, 1,804,366, with a call rate > 95%, a minimum allelic frequency (MAF) > 0.01, and a Hardy-Weinberg *P* value > 10^-6^ in controls were used for the two-step (phasing and genotype inference) imputation process. Haplotypes were phased with SHAPEIT2 v2.904 ^54^. For non-genotyped variants, genotype was inferred with IMPUTE2 ^55^, using the 1000 Genomes Phase III dataset as the reference panel and default values for all other parameters ^56, 57^. Only variants imputed with an *info* value ^55^ above 0.6 were retained for further analysis. In total, 10,014,109 autosomal variants with a MAF above 0.02 if genotyped (1,683,899 variants) or 0.05 if imputed (8,330,210 variants) were tested for association.

All variants for which evidence for association was obtained with a type I error of less than 5×10^−5^ (genotyped variants) or 10^-6^ (imputed variants) were retained for genotyping and association testing in the replication sample. As the replication sample was derived from exactly the same population as the discovery sample, we were able to prune all the significant variants on the basis of their estimated linkage disequilibrium (LD), to limit the genotyping effort. Among the variants of a given bin, defined as a group of variants with an r^2^ > 0.8 with a core variant, we first selected the genotyped variant with the highest likelihood of successful genotyping in the replication sample, according to the manufacturer’s instructions (see below). For bins without a genotyped variant or with genotyped variant for which the chances of genotyping success were low, we selected the imputed variant with the highest *info* value. Following LD pruning, 105 variants (99 genotyped and 6 imputed) were genotyped and tested for association with Buruli ulcer in the replication sample. These variants were genotyped by Illumina GoldenGate genotyping with VeraCode technology. The replication sample contained 708 individuals; 693 of these individuals (455 cases and 238 controls; Table 1) passed the quality control criteria (4 samples with a call rate < 95%, and 11 duplicated samples were excluded). Variants with a call rate < 95% or a Hardy Weinberg *P* value below 10^-4^ in controls were excluded.

Significant replication was defined as a variant having the same allelic effect as in the discovery sample, with a one-sided *P* value < 0.01 on the replication sample. As one of the replicated SNPs (rs9814705) was imputed in the discovery sample, we decided to perform Sanger sequencing on this SNP in the discovery sample to strengthen our conclusions. Sequences for the 803 samples were obtained with the Big Dye Terminator kit and a 3500xL automated sequencer from Applied Biosystems. Sequence files and chromatograms were inspected with GENALYS software ^58^. Given the high concordance between imputation and Sanger sequencing data (∼99%), genotypes inferred by the imputation process were used for the 143 missing Sanger-sequenced genotypes.

### Statistical analyses

Principal component (PC) analysis was performed on genotyped variants with a MAF > 0.05 to check for population structure, for the discovery sample and all the samples available from 1000 Genomes Project Phase III ^59^, with the *ad hoc* functions implemented in PLINK v1.9 ^60^.

Two statistical designs were considered for tests of the association between genetic variants and the occurrence of Buruli ulcer, depending on the phenotypic definition use. We first considered Buruli ulcer as a binary phenotype (affected/unaffected). A classical case/control design was used and all analyses were performed within a logistic regression framework. Gender had no significant effect (*P* value = 0.41) on binary Buruli ulcer status. Given that the controls were older than the cases by design, we did not include age as a covariate, to avoid over-adjustment, and the effect of age was accounted for by specific survival analysis methods (see below). We also evaluated the potential impact of cryptic population stratification and/or hidden relatedness within our discovery sample, by comparing the results of the association tests obtained with a classical logistic regression model and those obtained with a mixed-effect logistic model ^61^. We observed an extremely strong correlation (r = 0.90, Spearman *P* value < 10^-16^) between the top 1000 variants obtained by classical logistic regression and those obtained with a mixed model (Supplementary Fig. 2a). The largest difference for these top 1,000 variants, measured as the absolute difference between the –log10(*P* value) obtained in classical logistic regression that obtained with the mixed model, was less than 0.81. Consistent with these findings, we found no significant association between the three first principal components and the occurrence of Buruli ulcer.

We then considered age at onset for Buruli ulcer patients and age at examination for the exposed controls as the phenotype of interest. A survival analysis framework was used and all analyses were performed with Cox models ^62^. The analysis strategy was identical to that used for the binary phenotype: i.e., we evaluated the potential impact of cryptic population stratification and/or hidden relatedness within the discovery sample by comparing the results of the association tests performed with a classical Cox model with those obtained with a mixed-effect Cox model ^63^. Again, the correlation between the *P* values of the 1,000 top variants for the classical and mixed-effect Cox models was extremely strong (r = 0.97, Spearman *P* value < 10^-16^ (Supplementary Fig. 2a)). For these top 1,000 variants, the largest difference in terms of log10(*P* value) was at 0.27. Consistently, the use of the first three principal components had no impact on the results. The results reported here are therefore those for association tests performed with classical logistic or Cox models. All analyses were conducted assuming an additive genetic effect of the variants, and all the *P* values displayed were obtained in likelihood-ratio tests. All association analyses were performed with R (https://www.r-project.org/), PLINK 1.9 ^60^, GEMMA ^61^, coxme ^63^, SNPTEST v2.5.2 ^64^, and ProbABEL ^65^ software. Finally, we calculated the power of our discovery sample (402 cases and 401 controls) for detecting a significant association with Buruli ulcer coded as a binary trait, assuming a type I error of 5×10^−5^. Power estimates are shown for an additive model, by MAF, and effect size, as estimated by calculation of the odds ratio (OR) of the tested variants in Supplementary Fig. 5. For example, our sample had 50% power for the detection of variants with an OR of 1.6 and a MAF > 0.2, and 80% power for detecting variants with an OR of 1.8 and a MAF > 0.18. All these power estimates were calculated with QUANTO v1.2.4 (http://biostats.usc.edu/Quanto.html).

### Validation of variants previously found to be associated with common mycobacterial diseases

We used our discovery sample to assess genetic findings already reported for Buruli ulcer and in the larger context of common mycobacterial diseases. We recently identified a microdeletion in a familial form of Buruli ulcer that overlaps with a cluster of genes encoding defensins ^21^. We, therefore, first identified all the variants of the 50 defensin genes (located within 10 kb on either side of the gene) listed in the Ensembl GRCh37.p13 database and tested for association in our GWAS. We also considered all the variants reported to be eQTL (FDR < 0.05) for defensin genes in whole-blood and skin from the lower legs exposed to the sun in the GTEx database ^32^. Results were available for 5,452 defensin-related SNPs (Supplementary Table 1). We then searched for variants reported to be associated with Buruli ulcer in the three published candidate gene studies ^26, 27, 28^. A total of six variants on six genes were assessed. Finally, we investigated the role of host genetic factors potentially common to Buruli ulcer and the other two main mycobacterial diseases (i.e. tuberculosis and leprosy). We screened the GWAS catalog (https://www.ebi.ac.uk/gwas/) for variants associated with either tuberculosis or leprosy at the genome-wide level (*P* value < 5×10^−8^). We identified 36 independent SNPs, eight in TB, and 27 in leprosy, which we then assessed for association with Buruli ulcer in our GWAS discovery sample.

## Data Availability

The genome-wide variant genotyping and all other relevant data will be available upon request once the manuscript is published in a peer-reviewed journal.

## Acknowledgments

We thank Jean-Laurent Casanova, Emmanuelle Jouanguy and all the members of the HGID laboratory for useful discussions. We also thank Guillaume Vogt, Antoine Guérin, Jérémie Babonneau, Julien Guergnon, Grégoire Hure, Natasha Vladikine, and Myriam Berramdane for their contribution to genotyping and sequencing. We acknowledge support from the Fondation Raoul Follereau. LM and AA were supported by the Agence Nationale de la Recherche (ANR; grant no. ANR-17-BSV3-0013-01). JM, LA and AA were supported by the Laboratoire d’Excellence “Integrative Biology of Emerging Infectious Diseases” (grant no. ANR-10-LABX-62-IBEID) and the ANR under the “Investments for the Future” program (grant no. ANR-10-IAHU-01). QBV was supported by the Fondation Bettencourt Schueller through the MD/PhD program of the Imagine Institute. AA was supported by the Fondation pour la Recherche Médicale (grant no. DMI20091117308).

## Author Contributions

All authors were responsible for the study concept and design, and the analysis and interpretation of data. M-FA, CJ, LM, EM and AC collected the data. M-FA, LM, EM and AC did or confirmed diagnostics. QV, JM, MFA, EM, CJ, AC, MC, LL, IT and AA acquired the data. QV, JM, LA, and AA did the statistical analysis. JM, LA, and AA drafted the report. AA obtained the funding.

